# Evaluation of the performance of GPT-3.5 and GPT-4 on the Medical Final Examination

**DOI:** 10.1101/2023.06.04.23290939

**Authors:** Maciej Rosoł, Jakub S. Gąsior, Jonasz Łaba, Kacper Korzeniewski, Marcel Młyńczak

## Abstract

**Introduction:** The rapid progress in artificial intelligence, machine learning, and natural language processing has led to the emergence of increasingly sophisticated large language models (LLMs) enabling their use in various applications, including medicine and healthcare.

**Objectives:** The study aimed to evaluate the performance of two LLMs: ChatGPT (based on GPT-3.5) and GPT-4, on the Medical Final Examination (MFE).

**Methods:** The models were tested on three editions of the MFE from: Spring 2022, Autumn 2022, and Spring 2023 in two language versions – English and Polish. The accuracies of both models were compared and the relationships between the correctness of answers with the index of difficulty and discrimination power index were investigated.

**Results:** The study demonstrated that GPT-4 outperformed GPT-3.5 in all three examinations regardless of the language used. GPT-4 achieved mean accuracies of 80.7% for Polish and 79.6% for English, passing all MFE versions. GPT-3.5 had mean accuracies of 56.6% for Polish and 58.3% for English, passing 2 of 3 Polish versions and all 3 English versions of the test. GPT-4 score was lower than the average score of a medical student. There was a significant positive and negative correlation between the correctness of the answers and the index of difficulty and discrimination power index, respectively, for both models in all three exams.

**Conclusions:** These findings contribute to the growing body of literature on the utility of LLMs in medicine. They also suggest an increasing potential for the usage of LLMs in terms of medical education and decision-making support.

**What’s new?:** Recent advancements in artificial intelligence and natural language processing have resulted in the development of sophisticated large language models (LLMs). This study focused on the evaluation of the performance of two LLMs, ChatGPT (based on GPT-3.5) and GPT-4, on the Medical Final Examination across English and Polish versions from three editions. This study, to the best of our knowledge, presents the first validation of those models on the European-based medical final examinations. The GPT-4 outperformed GPT-3.5 in all exams, achieving mean accuracy of 80.7% (Polish) and 79.6% (English), while GPT-3.5 attained 56.6% (Polish) and 58.3% (English) respectively. However, GPT-4’s scores fell short of typical medical student performance. These findings contribute to understanding LLM’s utility in medicine and hint at their potential in medical education and decision-making support.

## 1. Introduction

The rapid advancements in artificial intelligence (AI), machine learning (ML) and natural language processing (NLP) methods have paved the way for the development of large language models (LLMs), which possess an unprecedented ability to understand and generate human-like texts. These models have demonstrated remarkable performance in various tasks, spanning from sentiment analysis, machine translation, to text summarization and question-answering [1], [2]. As a result, the potential application of LLMs in various domains, including medicine along with healthcare, is a topic of significant interest [3]. Recently, the AI topic has gained in even more general popularity thanks to the ChatGPT chatbot introduction for the public [4].

ChatGPT is a LLM developed by OpenAI and initially released on the 30^th^ of November 2022 on the website https://chat.openai.com/. ChatGPT became the fastest-growing application in history, as it gained 1 million users in 5 days and 100 million users just after 2 months after the initial launch. The first release of the service was based on the 3.5 version of the generative pre-trained transformer (GPT) model. The model was trained using Reinforcement Learning from Human Feedback technique with Proximal Policy Optimization (PPO) [5]. The training procedure contained three steps: (1) supervised learning where the AI trainer indicated the desired response, (2) training a reward model based on the ranking of different outputs, and finally (3) optimizing the policy against the reward model using the PPO. On the 14^th^ of March 2023, the newest version of the GPT model (GPT-4) was also released. Access to this model was restricted only to the premium users of the OpenAI chatbot (one can become a premium user only by taking out the subscription). GPT-3.5 and GPT-4 training data were cut off in September 2021, so those models were not exposed to the newest data. Both GPT-3.5 and GPT-4 performance were validated on the Massive Multitask Language Understanding (MMLU) test [6]. GPT-4 model outperformed other models not only in the English version but also after translation of the test to other languages (even those rarely used like Latvian, Welsh, or Swahili) [7].

In order to incorporate GPT-3.5/GPT-4 into a specific field it needs to be further validated in the field-specific tests. In medicine, the expertise of healthcare professionals is crucial in ensuring accurate diagnosis, effective treatment, and patients’ safety. To maintain a high standard of medical practice, rigorous assessment methods, such as different medical final examinations, are employed to evaluate the competency of medical graduates before they begin practicing independently. Such examinations cover a wide range of medical knowledge, including theoretical concepts, clinical reasoning, and practical skills, making it a suitable benchmark for evaluating the performance of LLMs in the medical domain [8], [9]. Results of validation analysis of GPT-3.5 on numerous medical examinations have been recently published [8], [10]–[15]. GPT-3.5 and GPT-4 were already validated on, to our knowledge, several national medical tests like the United States Medical Licensing Examination (USMLE) [9], Japanese [8], and Chinese National Medical Licensing Examinations [13], [14], on couple of medical benchmark databases like MedQA, PubMedQA, MedMCQA, and ABMOSS [15]– [17]. GPT-3.5 was also evaluated in terms of its usability in the decision-making process. Rao et al. reported that GPT-3.5 achieved over 88% accuracy by being validated using the questionnaire regarding the breast cancer screening procedure [18]. However, to the best of our knowledge, there have been no studies yet presenting the capabilities of GPT-3.5 and GPT-4 in terms of European-based medical final examinations.

In this paper, we hence aimed to investigate the utility of GPT-3.5 and GPT-4 in the context of Polish Medical Final Examination in two language versions – Polish and English. By evaluating the LLMs’ performance on the examination and comparing it to real medical graduates’ results, we seek to better understand their potential as a tool for medical education and clinical decision support as well as an improvement of the GPT technology which comes with the newest version of the model.

## 2. Methodology

Polish Medical Final Examination (Lekarski Egzamin Końcowy, or LEK, in Polish), which is necessary to complete medical education under Polish law and to pass to apply for the license to practice medicine in Poland (and based on the Directive 2005/36/EC of the European Parliament also in European Union). The exam is a test comprising 200 questions with 5 options to choose from and only a single correct answer. In order to pass a test, it is required to obtain at least 56% of correct answers [19]. As both models were trained on the data until September 2021 it was decided to evaluate their performance on 3 editions of the Polish Medical Final Examination – Spring 2022 (S22), Autumn 2022 (A22), and Spring 2023 (S23) in two versions – Polish and English. All questions from the previous editions of the examination are available online, along with the average results of medical graduates, detailing overall results, results of graduates who took the exam for the first time, those who graduated in the last 2 years, and those who graduated more than 2 years ago [20]. Besides the content of the question, the correct answers and answer statistics like the index of difficulty (ID) and discrimination power index (DPI) were published. Those indexes were calculated according to the equations presented below [21]:

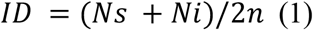

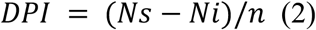

where *n* is the number of examinees in each of the extreme groups (27% of the participants with the best results and 27% with the worst results in the entire test), *Ns* – the number of correct answers to the analyzed task in the group with the best results, *Ni* – the number of correct answers for the analyzed task in the group with the worst results. The index of difficulty takes values from 0 to 1, where 0 means that the task is extremely difficult and 1 means that the task is extremely easy. The discrimination power index assumes values from -1 (for extremely badly discriminating tasks) to 1 (for extremely well discriminating tasks).

In the case of GPT-3.5, an API provided by OpenAI was used in order to accelerate the process of obtaining answers, as a model *gpt-3*.*5-turbo* was used [22]. From each response, the final answer was obtained and saved to the Excel file. If the answer was ambiguous, then the given question was treated as not answered (in other words – incorrectly answered). In the case of GPT-4, questions were taken as an input to the prompt of the models or with API using *gpt-4-0613* model. Final answers from all prompts were stored in Appendix 1 and 2 for GPT-3.5 and GPT-4 respectively.

The accuracy of both models for each test was calculated by dividing the number of correct answers by the number of all questions, which had the correct answer provided. As some questions were invalidated due to inconsistency with the latest knowledge, there were no correct answers for these questions, thus the number of correct answers was divided by the number smaller than 200. Questions which contained image were also excluded. Moreover, the Pearson correlation coefficient was calculated, and the Mann–Whitney U test was conducted to investigate the relationships between the correctness of the answers, the index of difficulty and the discrimination power index. The overall scores for each examination obtained by LLMs were also compared to the average score obtained by medical graduates who took the exam in the given editions. Consistency of responses depending on the language of the test was also validated by calculating the number of the same answers for each examination. All questions were asked between the 29^th^ of March and the 14^th^ of August 2023 (ChatGPT March 23 version). The significance level was set at the level of 0.05. For the usage of API, calculations, statistical inference, and visualizations Python 3.9.13 was used.

## 3. Results

GPT-3.5 managed to pass 2 out of 3 versions of examination in Polish and all 3 in English, while GPT-4 was able to pass all three versions of the exam regardless of language used. The detailed results obtained by both models are presented in Table 1 and visualized in Figure 1.

**Table 1.** Number of correct answers of GPT-3.5 and GPT-4 for each of the undertaken examinations. In the brackets, the number of questions with the given answers and percentage accuracy is provided next to the exam version and the number of correct answers respectively.

| Questions language | Model | S22 (195) | A22 (196) | S23 (194) |
| --- | --- | --- | --- | --- |
| Polish | GPT-3.5 | 98 (50.3%) – not passed | 113 (57.7%) – passed | 120 (61.9%) – passed |
|  | GPT-4 | 158 (81.0%) – passed | 161 (82.1%) – passed | 153 (78.9%) – passed |
| English | GPT-3.5 | 112 (57.4%) – passed | 117 (59.7%) – passed | 112 (57.7%) – passed |
|  | GPT-4 | 152 (77.9%) – passed | 159 (81.1%) – passed | 155 (79.9%) – passed |

**Figure 1.**
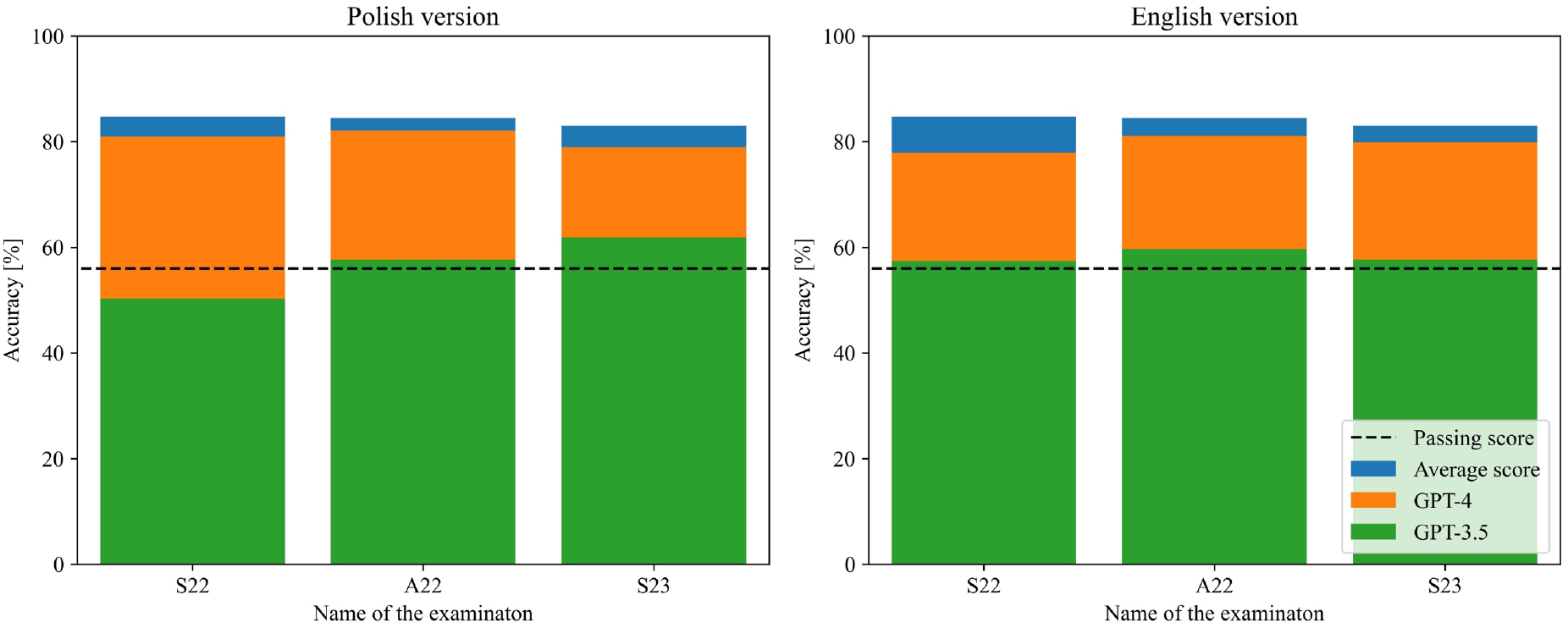
Comparison of the performance of both models along with passing score and average medical graduate score for all three examinations.

There was a significant positive correlation between the correctness of the answers and the index of difficulty as well significant difference between the index value for correct and incorrect answers in the case of all three exams for both models and languages. There was also a negative correlation and significant difference between the correctness of the answers and discrimination power index in the case of the A22 (for both languages) and S23 (only for Polish version) exams for both models. The results are presented in Table 2 and Table 3 for the index of difficulty and discrimination power index, respectively. The boxplots of the index values depending on the correctness of the answers were visualized in Figure 2 for the index of difficulty, and Figure 3 for the discrimination power index.

**Table 2.** Results of the correlation analysis with Pearson correlation coefficient and obtained p-value given in the brackets along with p-value obtained from the Mann–Whitney U test comparing the values of the index of difficulty for correct and incorrect answers.

|  |  |  | S22 | A22 | S23 |
| --- | --- | --- | --- | --- | --- |
| Polish | GPT-3.5 | Pearson correlation coefficient (p-value) | 0.319 (<0.001 ***) | 0.301 (<0.001 ***) | 0.174 (0.015 **) |
|  |  | P-value from Mann–Whitney U test | <0.001 *** | <0.001 *** | 0.004 ** |
| | GPT-4 | Pearson correlation coefficient (p-value) | 0.274<br>($<0.001$ ***) | 0.335<br>($<0.001$ ***) | 0.327<br>($<0.001$ ***) |
| | | P-value from Mann–Whitney U test | $<0.001$ *** | $<0.001$ *** | $<0.001$ *** |
| English | GPT-3.5 | Pearson correlation coefficient (p-value) | 0.229<br>(0.001 **) | 0.245<br>($<0.001$ ***) | 0.219<br>(0.002 **) |
| | | P-value from Mann–Whitney U test | $<0.001$ *** | $<0.001$ *** | 0.003 ** |
| | GPT-4 | Pearson correlation coefficient (p-value) | 0.430<br>($<0.001$ ***) | 0.273<br>($<0.001$ ***) | 0.307<br>($<0.001$ ***) |
| | | P-value from Mann–Whitney U test | $<0.001$ *** | 0.001 ** | $<0.001$ *** |

**Table 3.** Results of the correlation analysis with Pearson correlation coefficient and obtained p-value given in the brackets along with p-value obtained from the Mann–Whitney U test comparing the values of the discrimination power index for correct and incorrect answers.

|  |  |  |  |  |  |
| --- | --- | --- | --- | --- | --- |
|  |  |  | S22 | A22 | S23 |
| Polish | GPT-3.5 | Pearson correlation coefficient (p-value) | -0.092<br>(0.200 ns.) | -0.235<br>($<0.001$ ***) | -0.197<br>(0.006 **) |
|  |  | P-value from Mann–Whitney U test | 0.156 ns. | <0.001 *** | 0.009 ** |
|  | GPT-4 | Pearson correlation coefficient (p-value) | -0.111<br>(0.122 ns.) | -0.294<br>(<0.001 ***) | -0.208<br>(0.004 **) |
|  |  | P-value from Mann–Whitney U test | 0.098 ns. | <0.001 *** | 0.011 * |
| English | GPT-3.5 | Pearson correlation coefficient (p-value) | -0.098<br>(0.173 ns.) | -0.191<br>(0.007 **) | -0.109<br>(0.129 ns.) |
|  |  | P-value from Mann–Whitney U test | 0.116 ns. | 0.002 ** | 0.171 |
|  | GPT-4 | Pearson correlation coefficient (p-value) | -0.027<br>(0.705 ns.) | -0.151<br>(0.034 *) | -0.045<br>(0.555 ns.) |
|  |  | P-value from Mann–Whitney U test | 0.473 ns. | 0.055 ns. | 0.929 ns. |

**Figure 2.**
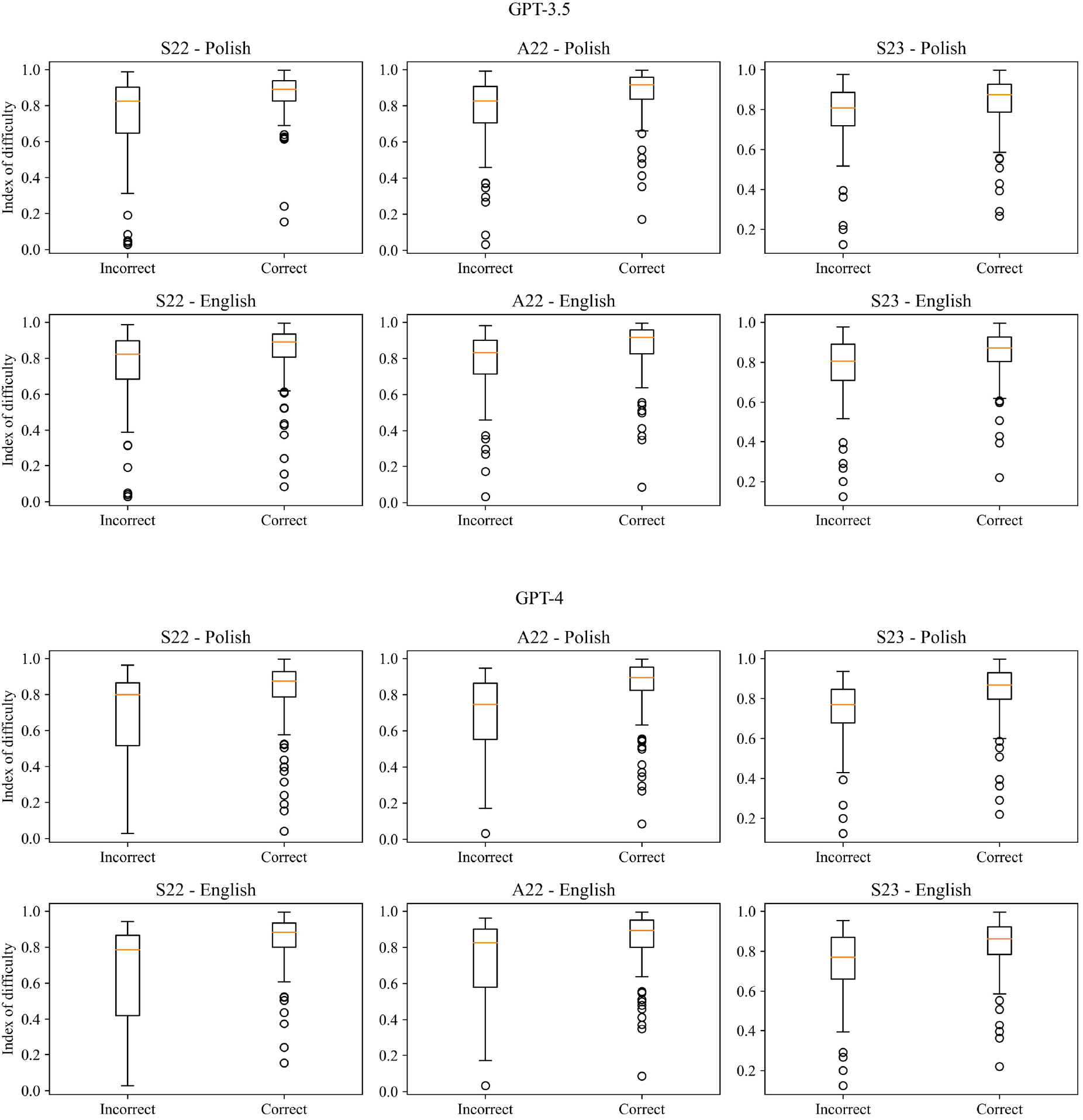
Boxplots of the index of difficulty for the correct and incorrect answers for all three versions of the examination and both languages.

**Figure 3.**
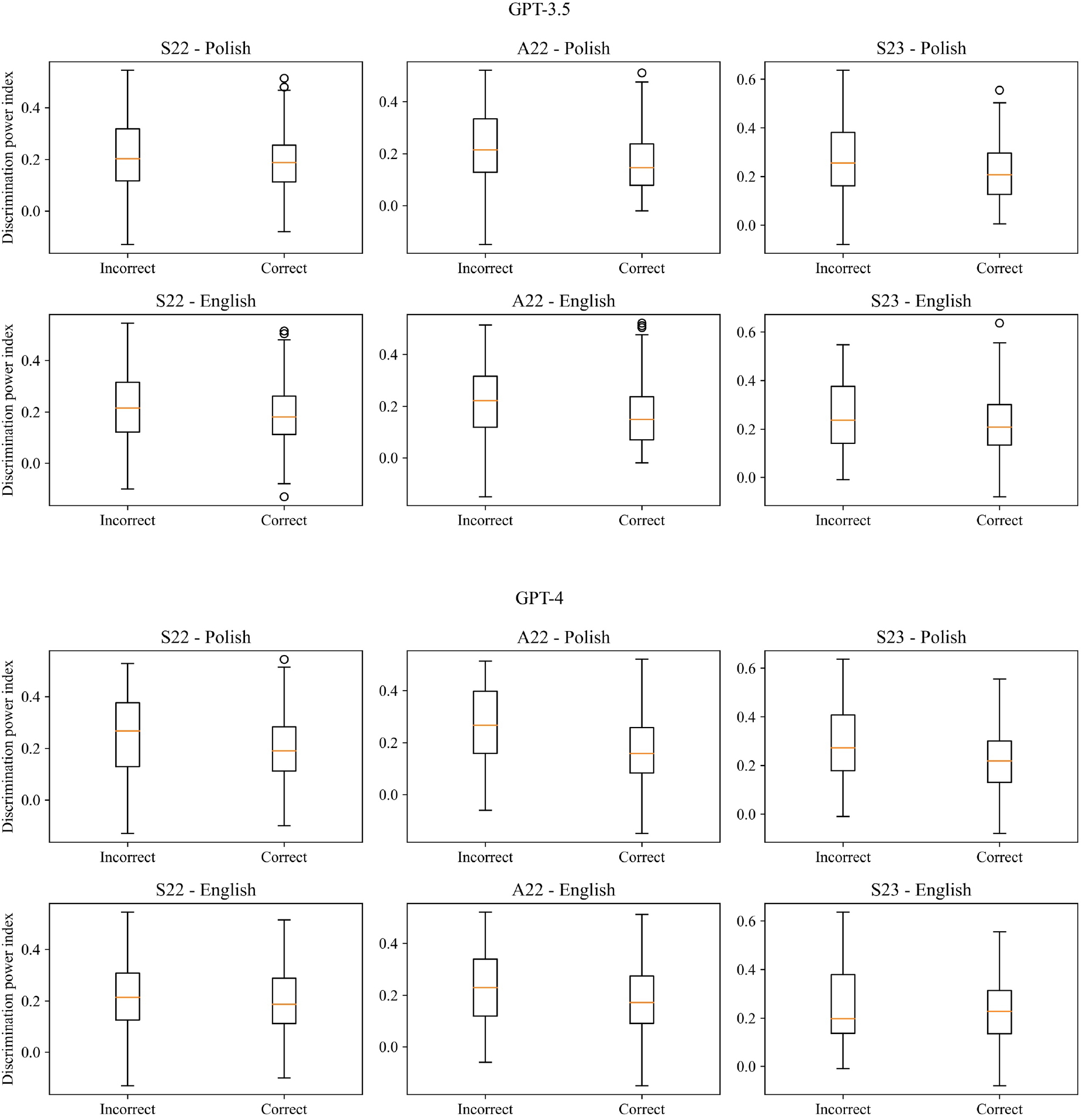
Boxplots of the discrimination power index for the correct and incorrect answers for all three versions of the examination and both languages.

The agreement between answers of the GPT models on the same questions in different languages is presented in Table 4.

**Table 4.** The number of questions on which models provided the same answer regardless of the test language. In brackets, the number of correct answers with the same response is presented.

|  |  |  |  |
| --- | --- | --- | --- |
|  | S22 | A22 | S23 |
| GPT-3.5 | 116 (79) | 113 (85) | 111 (86) |
| GPT-4 | 151 (134) | 161 (144) | 162 (143) |

## 4. Discussion

GPT-4 consistently outperformed GPT-3.5 in terms of the number of correct answers and accuracy across three Polish Medical Final Examinations. It indicates a significant improvement in the scope of medical knowledge represented by the GPT-4 model compared to the previous version. For both versions of the model, there is a significant correlation between the accuracy of the answers given and the difficulty of medical issues, indicating still a lack of in-depth knowledge in this area. Additionally, a negative correlation and significant difference were found between the correctness of the answers and the discrimination power index for both models in the A22 (both languages) and S23 (only Polish version) exams, which might be a sign of the simplicity of the model’s reasoning or the ability to simplify tasks in terms of the medical questions. In all versions of the test, GPT-4 scored slightly below medical student averages, which was equal to 84.8%, 84.5%, and 83.0% for S22, A22 and S23 respectively. The latest GPT version in the Polish test outperformed students who graduated over 2 years ago for S22 and those taking A22 as their first exam. Students who graduated less than 2 years before the examination consistently outperformed both GPT models in both languages. The consistency of the answers between different language versions of the test was much higher for GPT-4 than for GPT-3.5. On average, the most recent model returned identical answers across test languages in 81.0% of instances, compared to GPT-3.5’s 58.1% consistency. This highlights the improvement of text understanding and knowledge of the GPT-4 model. On average, GPT-3.5 exhibited a 1.6% higher accuracy in answering English questions than Polish ones. On the contrary, GPT-4 showed a 1.0% higher accuracy in Polish over English, which contrasts with the evaluation on the Massive Multitask Language Understanding (MMLU) benchmark, where accuracy in Polish was 3.4% lower than in English [7].

Our results on the European-based medical final examination are in line with other studies conducted on different tests and languages from North America and Asia, which indicated the improvement of the medical knowledge possessed by GPT LLMs alongside with the development of the consecutive versions. Kung et al. evaluated the performance of GPT-3.5 on the USMLE, where GPT-3.5 outperformed its predecessor (GPT-3) with a score near or passing the threshold of 60% accuracy, which is required to pass the exam [9]. Recently, GPT-4 model was also evaluated on USMLE by Nora et al. The newest version of the GPT model outperformed GPT-3.5 with the improvement of its accuracy by over 30 percentage points [16]. In this study, GPT-4 turned out to be superior compared to its previous version and Flan-PaLM 540B model [17] in the evaluation on other medical benchmarks like MedQA, PubMedQA and MedMCQA. In the study performed by Gilson et al., GPT-3.5 was confronted with the commonly used ABMOSS medical question database and 120 free questions from the National Board of Medical Examiners (NBME) [15]. GPT-3.5 outperformed IntructGPT and GPT-3 models in terms of accuracy by at least 4.9% and 24%, respectively. As shown by Kasai et al., GPT-4 was also able to pass the Japanese Medical Licensing Examinations again outperforming GPT-3.5 [8]. This study also highlighted the relationship between the correctness of the answers given by the LLM and the difficulty of the questions, which was also reported in our results. Study performed by Mihalache et al. presented that GPT-3.5 performed the best in the general medicine questions, while obtaining the worst results in the specialized questions [23]. Bhayana et al. demonstrated that GPT-3.5 exhibited superior performance on questions that required low-level thinking compared to those which require high-level thinking [24].

Moreover, the model struggled with questions involving the description of imaging findings, calculation and classification, and applying concepts. Recently, Google and DeepMind presented their LLM PaLM 2 and its medical domain-specific finetuned MedPaLM 2 [25], [26]. The performance of GPT-4 and MedPaLM 2 on USMLE, PubMedQA, MedMCQA and MMLU appears to be very similar, where both GPT-4 and MedPaLM 2 were superior to each other in an equal number of tests evaluated. In this comparison, it is worth noticing that GPT-4 is a general-purpose model and was not explicitly finetuned for the medical domain.

There may be several potential reasons for the imperfect performance and providing incorrect answers by the tested models. First of all, both models are general-purpose LLMs that are capable of answering questions from various fields and are not dedicated to medical applications. This problem can be addressed by fine-tuning the models, that is, further training them in terms of medical education. As was shown in other studies, a finetuning of LLMs can further increase the accuracy in terms of answering medical questions [27]–[29]. Currently, OpenAI does not provide finetuning options for GPT-3.5 and GPT-4, but in the future, when this feature becomes available, it is also planned to explore the capabilities of those models after the finetuning on a medical dataset. In order to further increase the model’s accuracy in terms of medical questions the medical databases should be expanded, and instruction prompt tuning techniques could be applied [17].

We believe that the appearance of such powerful tools might have a significant impact on the shape of the public health and medicine of tomorrow [30]. ChatGPT already offered evidence-based advices to public health questions from addiction, interpersonal violence, mental health, and physical health categories [31]. Accurate and validated LLMs with broad medical knowledge can be beneficial for medical students in terms of self-learning, e.g., by generating tailored learning materials, improving physician-patient communication by simulating conversations and clinical reasoning by providing step-by-step explanations of medical cases [32]. This influence will not be restricted to education, but also it might be useful in terms of taking a medical note from a transcript, summarization of test results, or decision-making support [4], [32]–[36]. Moreover, LLMs could also be useful for the personal assistants’ solutions and provide reasonable recommendations in the field of public health e.g., quitting smoking [31]. The importance of prompt engineering (the way of asking questions) should also be emphasized because it affects the quality of the generated answers [37], [38]. Also, a recent study has shown that chatbot responses were preferred over physician responses on a social media forum, which shows that AI may significantly improve the quality of medical assistance provided online [39]. However, it is also important to check the authenticity of the responses generated by GPT model, as it might “hallucinate”, especially regarding provided references [40]–[42]. Alongside other researchers, we believe that LLMs although they need to be approached with caution, are not a threat to physicians [43], but can be a valuable tool and will be used more widely in the near future [4], [44], [45]. As of now, it is necessary to remember, that still a human should be at the end of the processing chain.

While the results of this study demonstrated the potential utility of AI language models in the medical field, several limitations should be acknowledged. First of all, the study focused solely on the Polish Final Medical Examination, which may limit the generalizability of the findings to other medical examinations or languages. What is more, PFME is an A-E test, which means that in some cases the correct answers could be by chance not as the result of the knowledge possessed by the models. Moreover, although GPT-4 outperformed GPT-3.5, the overall accuracy of both models was still suboptimal and worse than the average for medical students. This emphasizes the need for further improvements in LLMs before they can be reliably deployed in medical settings e.g., for self-learning or decision-making support.

In conclusion, this study highlights the advances in AI language models’ performance on medical examinations, with GPT-4 demonstrating superior performance compared to GPT-3.5 regardless of the language used. However, there is still considerable room for improvement in their overall accuracy. Future research should focus on finetuning of those models and exploring their potential applications in various medical fields, such as diagnostic assistance, clinical decision support, and medical education. Further tests of LLMs could also include more open questions with evaluation by physicians without prior knowledge of the origins of the answers (if it was created by LLM or a human being).

## Data Availability

All data produced in the present study are available upon reasonable request to the authors

https://cem.edu.pl/

## 5. Author Contributions

Conceptualization: Maciej Rosoł

Data Collection: Maciej Rosoł, Kacper Korzeniewski, Jonasz Łaba

Data Curation: Maciej Rosoł

Formal Analysis: Maciej Rosoł

Methodology: Maciej Rosoł

Visualization: Maciej Rosoł

Writing – Original Draft Preparation: Maciej Rosoł

Writing – Review & Editing: Maciej Rosoł, Jakub S. Gąsior, Marcel Młyńczak, Kacper Korzeniewski, Jonasz Łaba

